# General population screening for type 1 diabetes using islet autoantibodies at the preschool vaccination visit: a proof-of-concept study (the T1Early study)

**DOI:** 10.1101/2023.11.03.23297978

**Authors:** Claire Scudder, Julia Townson, Kathleen M Gillespie, Jane Bowen-Morris, Philip Evans, Sarah Jones, Nicholas P B Thomas, Jane Stanford, Robin Fox, John A Todd, Sheila Greenfield, Colin M Dayan, Rachel E J Besser

## Abstract

**Objective:** Type 1 diabetes (T1D) screening programmes testing islet autoantibodies (IAbs) in childhood can reduce life-threatening diabetic ketoacidosis. General population screening is required to detect the majority of children with T1D, since in >85% there is no family history. Age 3-5 has been proposed as an optimal age for a single screen approach.

**Design:** Capillary samples were collected from children attending their pre-school vaccination and analysed for IAbs to insulin, glutamic acid decarboxylase, islet antigen-2, and zinc transporter 8, using Radiobinding/Luciferase Immunoprecipitation Systems assays. Acceptability was assessed using semi-structured interviews and open-ended postcard questionnaires with parents.

**Setting:** Two primary care practices in Oxfordshire, UK.

**Main outcome measures:** The ability to collect capillary blood to test IAbs in children at the routine pre-school vaccination (3.5-4 years).

**Results:** Of 134 parents invited, 66 (49%) were recruited (median age 3.5y (IQR 3. 4-3.6), 26(39.4%) male); 63 provided a sample (97% successfully). Parents (n=15 interviews, n=29 postcards) were uniformly positive about screening aligned to vaccination and stated they would have been less likely to take part had screening been a separate visit. Themes identified included preparedness for T1D, and the long-term benefit outweighing short-term upset. The perceived volume of the capillary sample was a potential concern and needs optimising.

**Conclusions:** Capillary IAb testing is a possible method to screen children for T1D. Aligning collection to the pre-school vaccination visit can be convenient for families and allows a universal approach without the need for an additional visit.

*KEY MESSAGES:* What is already known on this topic?
Screening children for type 1 diabetes by measuring islet autoantibodies (IAbs) may reduce life-threatening diabetic ketoacidosis. The optimal age for screening children at a single timepoint has been proposed as age 3-5. Routine immunisations are given at a similar age. What does this study add?
Aligning IAb testing with the pre-school vaccination visit (age 3.5-4y) is feasible and acceptable. Potential barriers and facilitators of this approach are explored. How this study might affect research, practice or policy?
The routine vaccination programme is a potential opportunity to screen children for future type 1 diabetes, offering improved engagement and potentially reducing the costs associated with a screening programme; all of which need exploration in a large and definitive study.

## INTRODUCTION

Type 1 diabetes (T1D) is characterised by well-defined stages, with a preclinical phase preceding clinical symptoms (1). The presence of two or more islet autoantibodies (IAbs) to insulin (IAA), glutamic acid decarboxylase (GADA), islet antigen-2 (IA-2A), and zinc transporter 8 (ZnT8A) confers >80% risk of children progressing to insulin requirement during childhood (2).

In Europe and North America, 15-70% of children present with diabetic ketoacidosis (DKA) at diagnosis (3, 4). This typically requires hospitalization, and causes psychological and physiological morbidity (4–6). Screening programmes measuring IAbs significantly reduce rates of DKA at diagnosis (7, 8), hospitalization (9), and the presenting level of glycaemia (10, 11).

Adopting population screening for T1D requires a universal approach, since the majority (>85%) of children do not have a relative affected. It has been suggested that the optimal age to test for IAbs at a single time point is between 3 to 5 years (8, 10, 11), since peak IAb development occurs between 9 months and 2 years (2). Our aim was to explore, as a ‘proof-of-concept’ study, general population screening for IAbs, in children attending their routine pre-school vaccination in primary care. In this context, we define a ‘proof of concept’ study, as informing any ethical implications and providing the foundations of knowledge (12), rather than assessing effectiveness.

The primary outcome was the uptake of capillary IAb testing in children attending their pre-school vaccination. Secondary outcomes included (1) blood and serum volumes collected; (2) the ability to measure all four IAb (IAA, GADA, IA2A, ZnT8A); (3) acceptability from parents whose children were screened, and (4) feedback from non-responders.

## METHODS

### Participants

Between June 22^nd^ and November 29^th^ 2022, across two primary care practices (PCP) in Oxfordshire, UK, the parents of children, aged 3.5 - 5 years, scheduled for their pre-school vaccination (Diphtheria, Tetanus, Pertussis, Polio, and MMR), were invited to participate (Figure 1). Invitations were sent by text message attached to an automated pre-school vaccination invitation and followed up by telephone.

**Figure 1.**
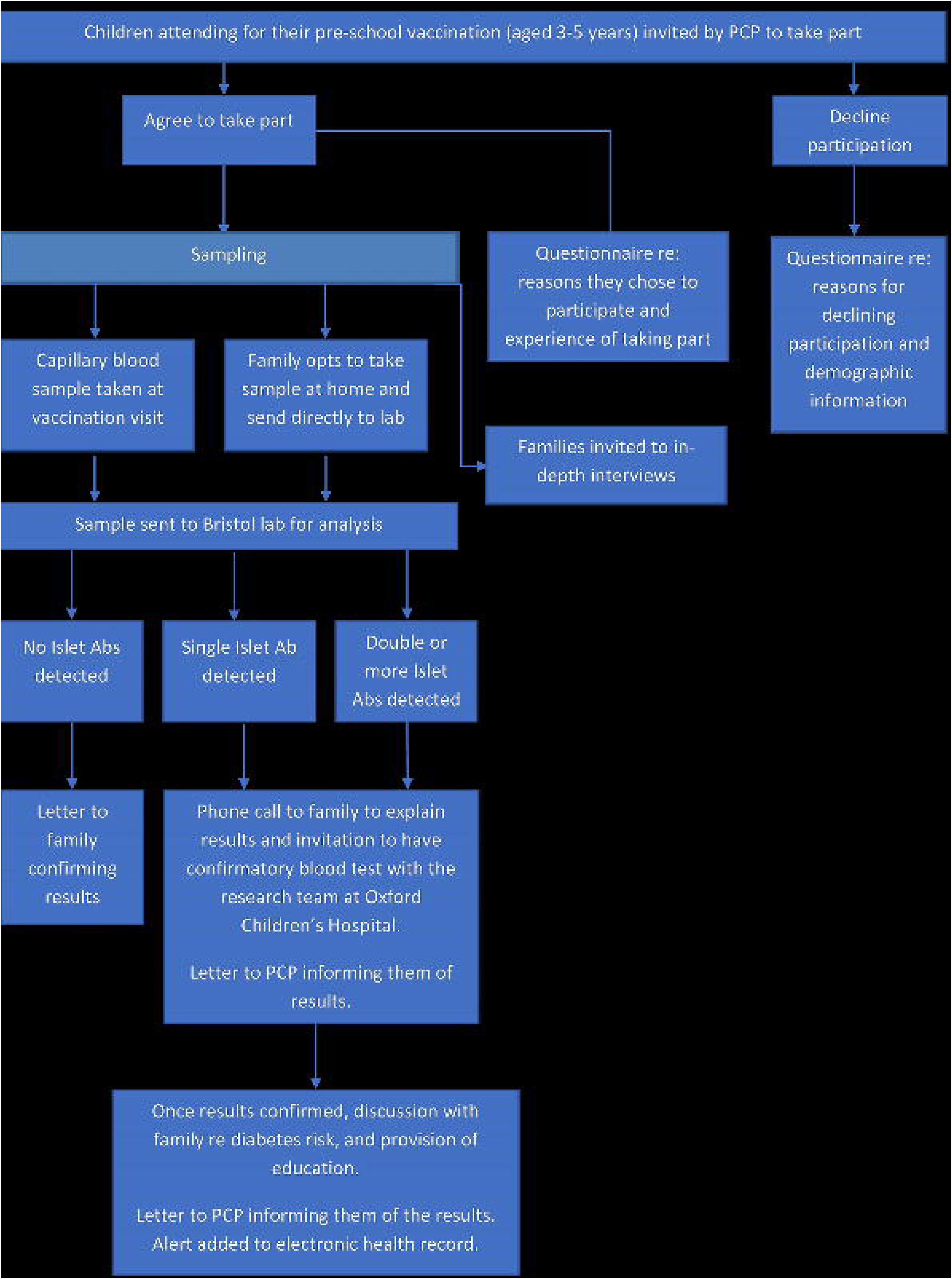
Study Schema.

Our approach was pragmatic and exploratory, to align with each PCP. This allowed each PCP to decide on their recruitment method; either sending invitation texts to parents of children due vaccination, or to those who had already booked a vaccination (Supplemental Table 1: eligibility criteria). Informed consent was obtained from parents in-person or remotely, prior to screening (Supplemental Methods 1: participant information sheet).

### Sampling and Laboratory methods

During or immediately after vaccination, research nurses collected a capillary sample up to 200 µl (Supplemental Methods 2: blood collection) and posted it to the University of Bristol Alistair Williams Antibody Facility (AWAF), UK. Samples were processed for serum isolation on arrival. IAA, GADA, IA-2A and ZnT8A were measured by Radiobinding assays (RBA)(13) for serum volumes >60 µL, and by Luciferase Immunoprecipitation Systems (LIPS) assay if <60 µL (14).

### Qualitative methods

#### i. Interviews

Participants were approached by email, after their child’s participation in screening, using a convenience sampling strategy. Fifty-one parents were approached, and 15 took part (all mothers), 2 declined, and 34 did not provide a response. Interviews were planned prior to receiving the test results. However, five participants had already received the results at the time of interview. Participants were remunerated for their time (£25 gift voucher).

### Data collection

The interview schedule (Supplemental Table 2) assessed the acceptability of the screening process. Interviews were conducted by CS, online, via Microsoft Teams or telephone, depending upon the participant’s preference, with oversight and training from JT, recorded verbatim and transcribed.

#### ii. Postcards

Parents (n=66), and those declining participation, were provided with postcards containing up to six questions to complete anonymously immediately after the vaccination/screening appointment (Supplemental Table 3), and which broadly mirrored the interview schedule. Postcards provide an opportunity to collect data with minimum burden to the participants whilst reducing the potential for recall, outcome, and emotional bias (15).

### Analyses

The data was analysed inductively and deductively (16). An established and defined process for developing a codebook in thematic analysis was followed (17). CS and JT each applied initial codes to the raw, redacted data from the first three participant interviews, using NVivo 12. These initial codes identified by both researchers were discussed, with additional members of the study team (RB, consultant paediatric endocrinologist) and SG, medical sociologist. From this discussion a coding framework was developed, using hierarchical themes, sub themes, and a description of the meaning of each code. A deductive approach was then applied to the coding of all data, including the postcard data, using the coding framework. Following coding by both researchers, consensus relating to specific themes was reached with the larger group). Finally, identification and interpretation of the emerging themes was discussed and agreed (JT, CS, SG, RB).

### Data collection

Demographic data were collected to allow assessment of deprivation score, using the Index of Multiple Deprivation (IMD), where 1 is most deprived and 10 least deprived.

### Ethical approval

Northwest – Preston Research Ethics Committee (ref 21/NW/0340)

## RESULTS

### Population

Of 134 eligible children identified and their respective parents invited to participate, 66 (49%) were recruited, see Figure 2. PCP 1 invited families that had already booked their child’s vaccinations, and PCP 2 invited families whose children were due vaccination. More families responded and were recruited with the former approach (93% vs 45% response rate, 88% vs 62% recruitment). Of the 88 parents who responded, 9 (10%) declined the study, 4 (5%) refused vaccination, and 9 (10%) were not recruited for other reasons (no reason provided (n=5), appointment availability (n=3), did not attend (n=1)).

**Figure 2.**
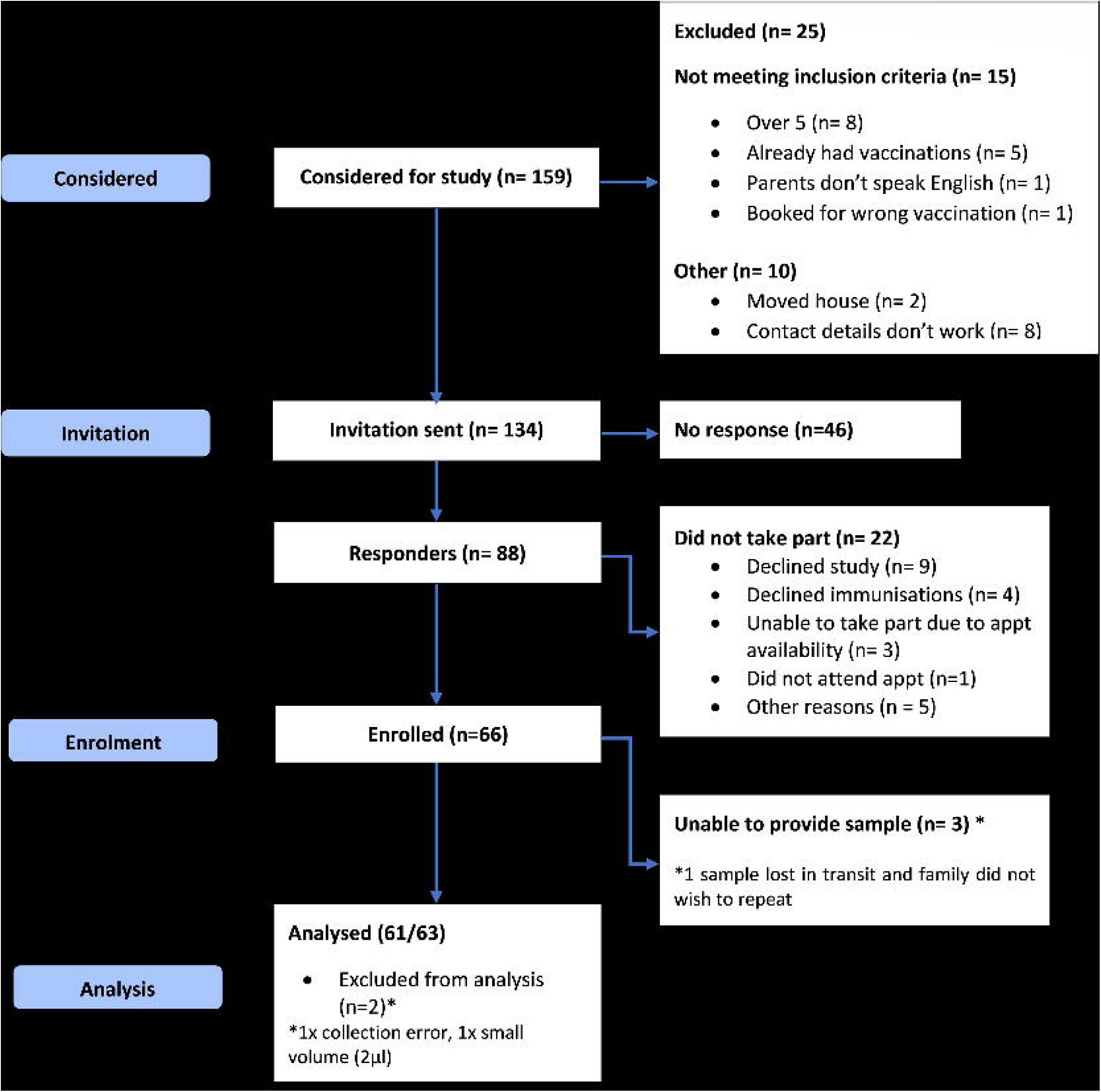
CONSORT diagram. Consolidated Standards of Reporting Trials

The 66 children recruited had a median age 3.5y (IQR 3.4-3.6, range 3.1-5.1y), 40 (60.1% female) and in the majority (95%) there was no family history of T1D. The study population was predominantly white British (75.8%) and moderate to high affluence (IMD median decile 8 (IQR 5-9, range 3-10)).

### Capillary islet autoantibody collection

Of the 66 participants enrolled, 63 (95%) provided a sample, two did not provide a sample, one provided a sample but was lost in transportation. Two samples were collected at home..

There was a 95% (60/63) success in sampling all four IAbs; one participant had a low serum volume resulting in only GADA, IA-2A and ZnT8 being analysed, and two samples failed, one due to low volume and the other due to collection error. The median serum volume collected was 100 µL (80–155), with 83% (52/63) >60 µL; Table 1.

**Table 1.** Participant demographics (n=66) and baseline data from samples available for analysis (n=61).

|  |  |
| --- | --- |
| **Age median (IQR) years** | 3.5(3.4-3.6) |
| **Female sex, n (%)** | 40(61) |
| **Ethnicity, n (%)** |  |
| English/Welsh/Scottish/Northern Irish | 50 (75.8) |
| Other white background | 4 (6.1) |
| Chinese | 3 (4.5) |
| Indian | 3 (4.5) |
| Gypsy or Irish traveller | 2 (3.0) |
| White & Asian | 1 (1.5) |
| Other Mixed/multiple ethic background | 1 (1.5) |
| Other ethnic group | 1 (1.5) |
| Not stated | 1 (1.5) |
| **Family history of type 1 diabetes, n (%)** | 3(4.8) |
| **IMD\* decile, median (IQR)** | 8(5-9) |
| **Whole blood volume, median (IQR) µL** | 200(150-220) |
| **Serum volume, median (IQR) µL** | 100(80-155) |
| **Time to results, median (IQR) days** | 29 (25-46) |

*\*Index of Multiple Deprivation (IMD) ranks small areas in England from 1 (most deprived) to 32,844 (least deprived) and divides into 10 equal groups: most deprived (1) to the least deprived (10).*

One participant was screened and confirmed positive by RBA for IAA. The family attended an appointment with the paediatric diabetes team to receive T1D education and were offered repeat islet autoantibody testing after six months. They also underwent random glucose and HbA1c measurement, which were normal. The median time to receive results was 29 days (25–46).

### Qualitative study results

15 parents participated in the qualitative sub-study, interviews were 17 - 32 minutes long. Thirty-two postcards were returned, 13 from participants attending PCP 1, and 16 from PCP 2; Supplementary Table 4.

Three hierarchical themes were identified, Table 2.

**Table 2.**
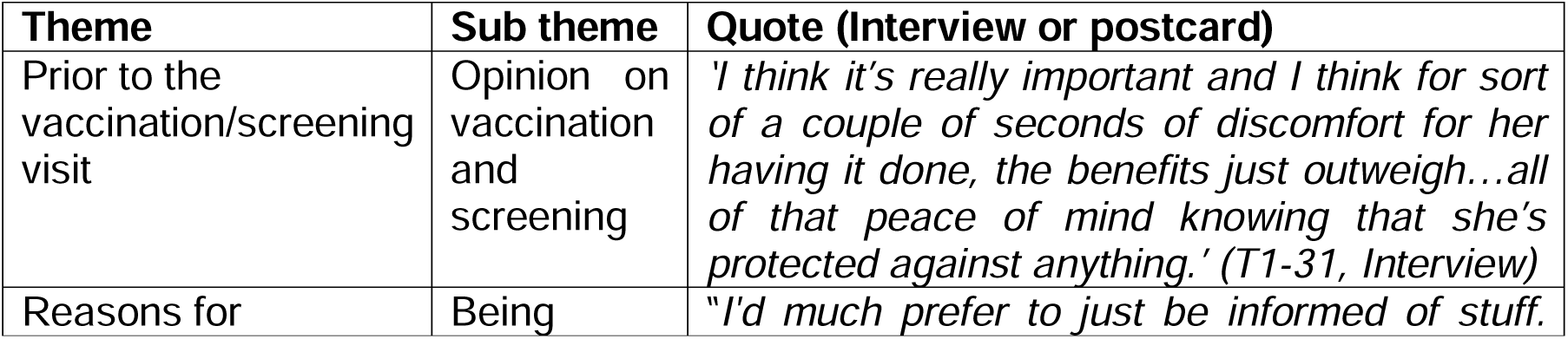
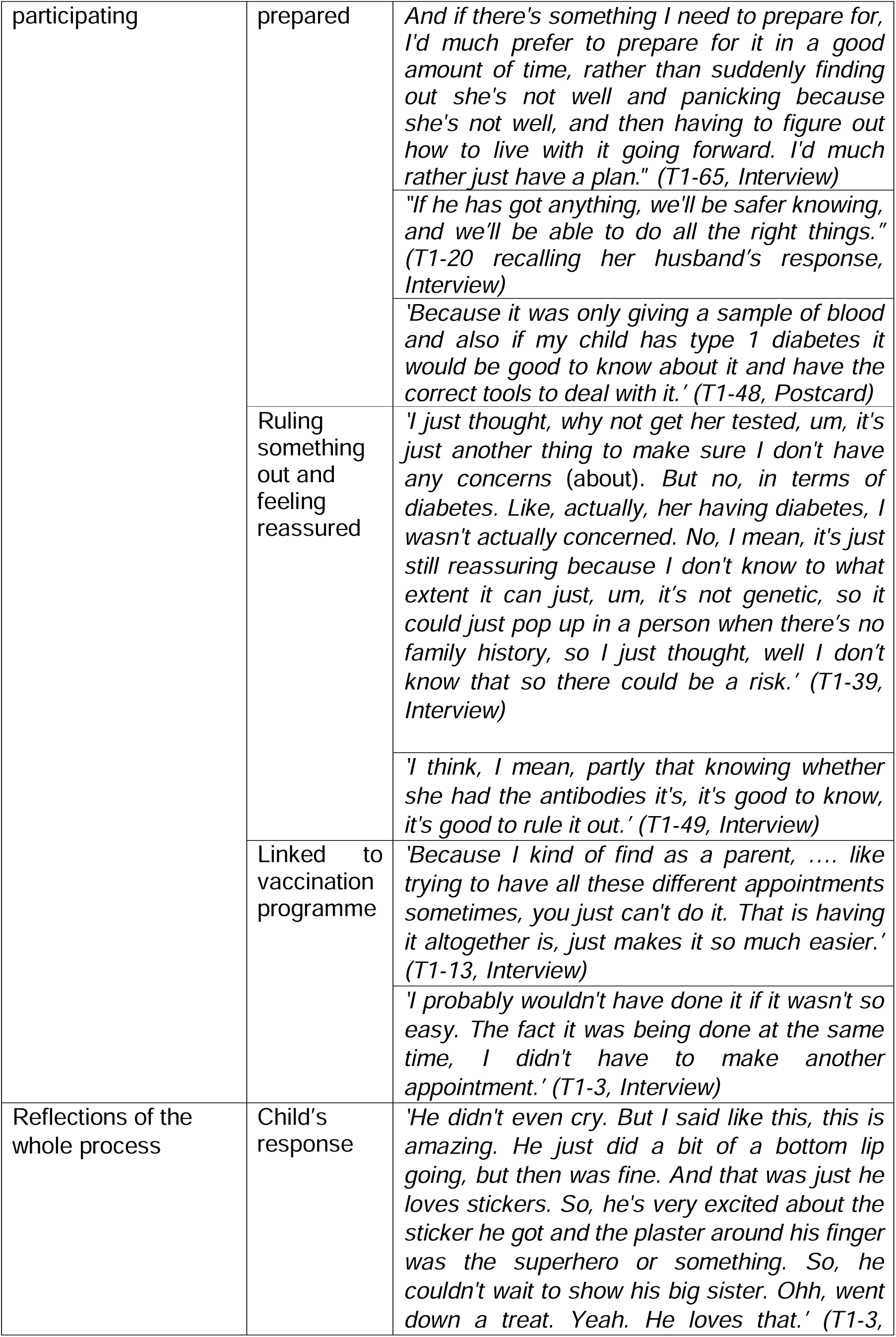
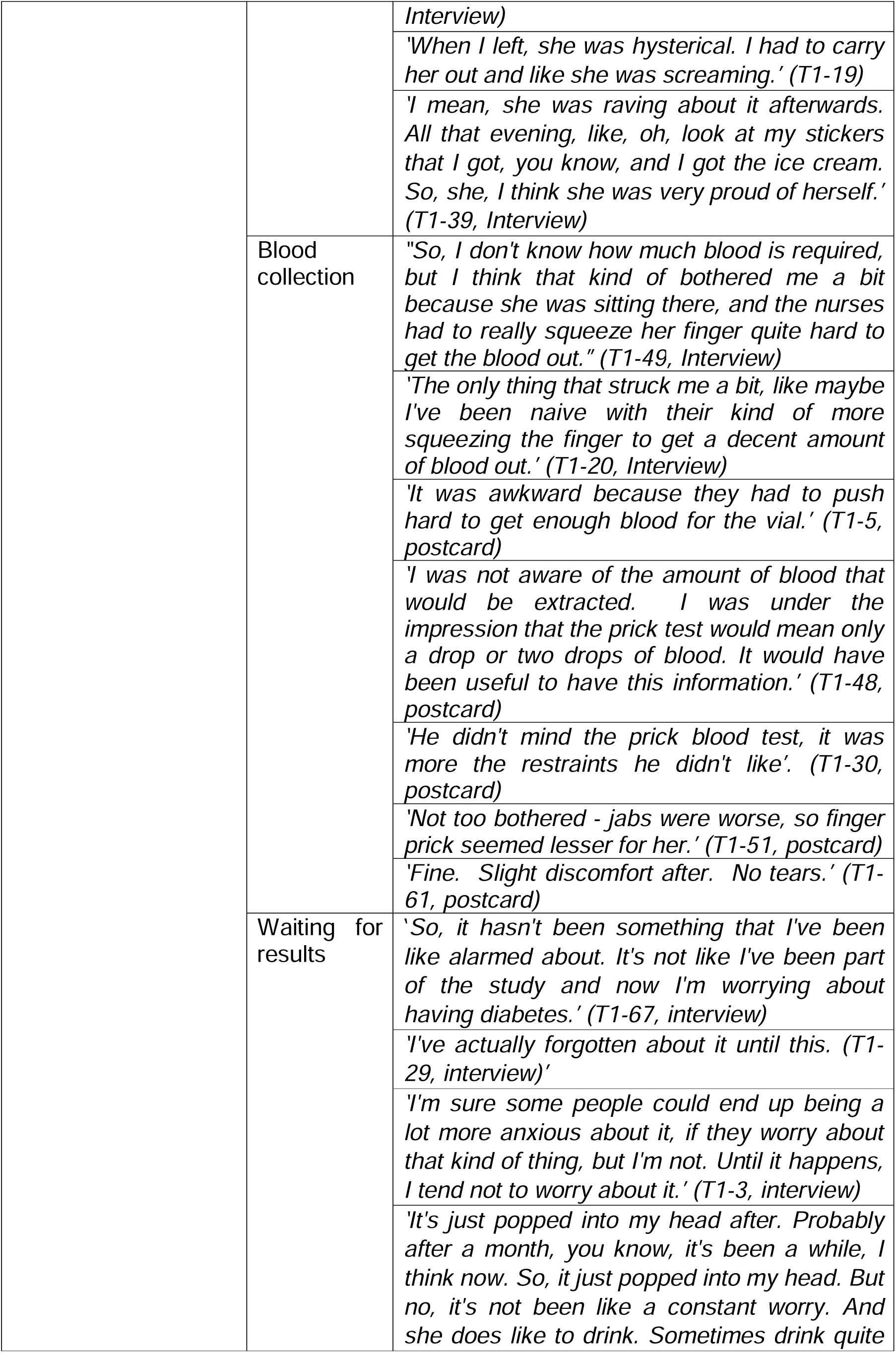
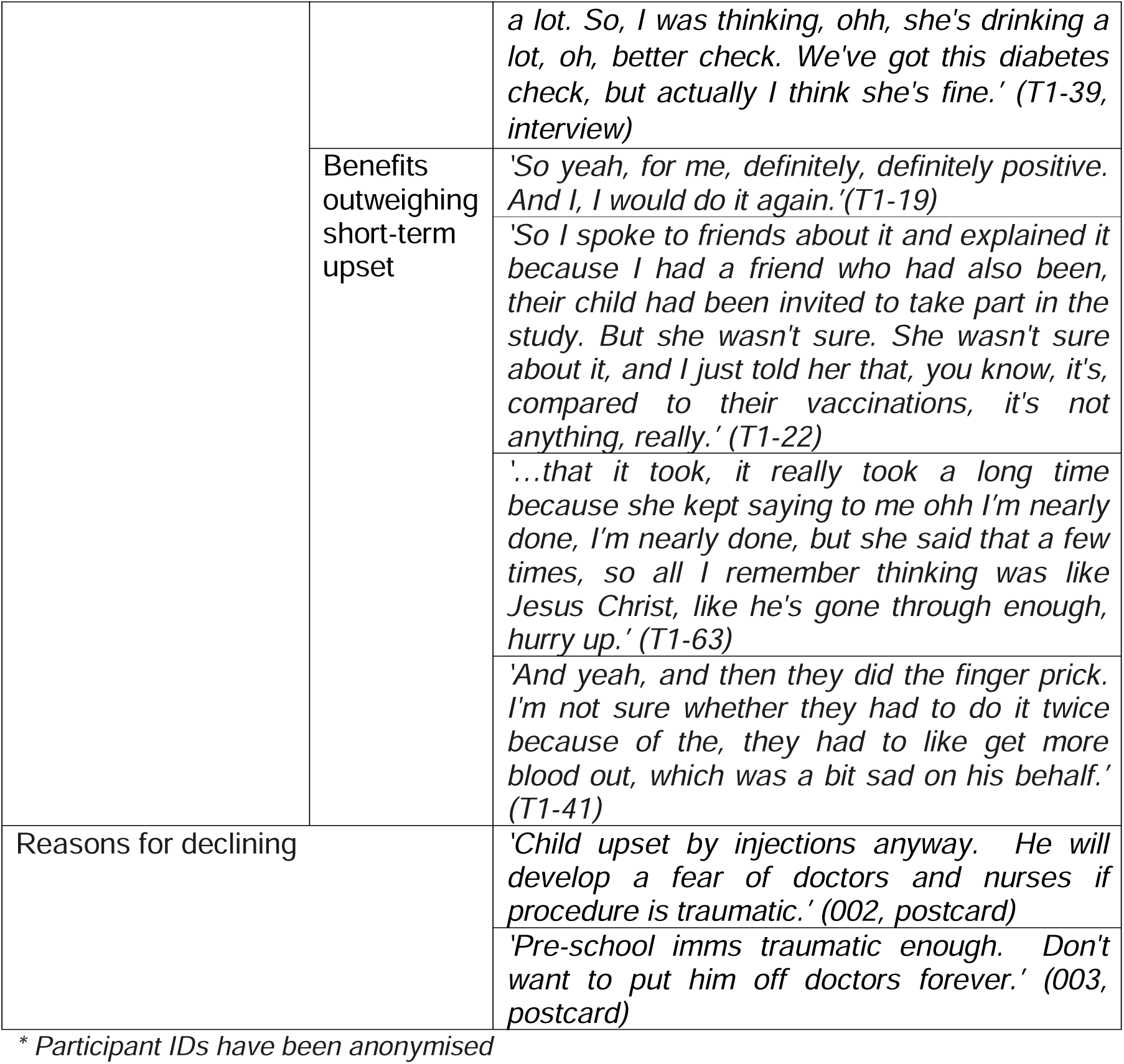
Participant quotations from the data against the named themes and subthemes*.

### Prior to the vaccination/screening visit

All parents recognised the importance of childhood vaccination to protect against childhood illnesses. This positive attitude was reflected when considering undergoing screening for T1D. Parents described the benefits of knowing about risks to their child’s health.

### Reasons for participating

#### Being prepared

Overwhelmingly parents recalled they would rather be prepared, allowing them to adjust to a potential diagnosis of T1D. Having time to mentally prepare, gather information and plan ahead was considered preferable to having to deal with the shock of a sudden diagnosis. Some parents related their opinions to their family background or general views on managing the health of their family.

#### Ruling something out and feeling reassured

Participants described wanting to know the outcome, stating that having the results provided reassurance.

#### Linked to the vaccination programme

The majority of parents reflected positively on having screening aligned to the child’s vaccination programme, and the fact that it was conducted at the PCP surgery. Parents recounted busy lives, lack of transport, having to take their child out of nursery, as reasons why they preferred the screening test to be conducted as part of a routine health visit. Some reflected they would have been less likely to agree to take part had the screening test been offered separately or in a different location.

### Reflections on the whole process

#### Child’s response

Parents described mixed experiences of their child’s response to the process. Those reflecting positively described the ease of the process. Negative experiences included having to hold their child securely, and the stress caused to their child. Most parents reflected that their child was primarily upset due to the vaccinations, which were carried out first. Most parents stated that their child had recovered after 5 to 10 minutes.

Some parents were surprised about how well their child had reacted and that their child had not mentioned it afterwards, whilst others recalled their child mentioning their finger or arm hurting.

One parent related that they had been worried that the screening process might give their child a lasting fear of going to the doctors but was reassured that their child still seemed keen to attend afterwards.

In general, parents recalled their child’s excitement over the stickers, plaster or treat they had received because of the visit.

#### Blood collection

Parents were surprised by the amount of blood required, and how much pressure the nurse needed to apply to their child’s finger to collect the blood. There was concern over the length of time it took to collect the volume of blood required. Parents recounted feeling distressed and wishing the nurse would finish, and feeling sad on behalf of their child.

#### Waiting for results

In the main, parents had either forgotten about the results or expressed that they were not worried about them. Two parents recounted being nervous about getting the results, and one parent recalled attributing their child’s behaviour as a potential indicator of the symptoms of T1D, acting as a reminder to check their results.

#### Benefits outweighing short term upset

Overall, parents recalled satisfaction with the process, describing that the benefits outweighed the potential upset.

Parents were glad they had taken part and although, in some cases, the experience caused their child distress, overall, they felt positive about the experience and related that they would do it again and recommend it to others. All parents stated that they would want their child to take part in screening for T1D again if asked.

### Postcard data from individuals who declined

Three postcards were returned from parents who had declined participation, and all went ahead with vaccinations. Two of the three parents stated that they felt undergoing the immunisations was difficult for their child and they did not want them to develop a fear of doctors and nurses. The other participant recounted they were not able to consider the information at that time.

## DISCUSSION

In this proof-of-concept study, we explore the methodology of collecting capillary blood for IAbs in children at the time of the pre-school vaccination visit. We report that 49% participants were recruited, although this was higher (88%) in parents who were approached after they had booked their child’s vaccination visit. Once the child attended, the success rate of measuring all four IAbs was high (95%). Although the data is limited and exploratory, we found that the experience of IAb testing was broadly positive.

### T1D screening at the pre-school vaccination versus other approaches

A single screen IAb test has been suggested between ages 3-5 years (18, 19) with sensitivity around 40%. This approach was taken in the Fr1da study which initially screened children between 1.75 and 6 years, during the ‘well child check’ in primary care (7). No such routine health visit exists in the UK but the pre-school vaccination at age 3.5-5 years offers a similar opportunity.

Of the 88 parents who responded, only nine actively declined participation. It is unclear whether the lack of response from individuals who were initially contacted via text messaging (n=46) was due to a lack of interest in the study or other reasons, such as not reading the information leaflet. A similarly low study uptake in response to primary care text messaging has been reported, and should be considered when expanding recruitment (20).

Several research groups are now testing non-targeted, opportunistic screening (4, 21). However, universal screening would allow population DKA reduction, in accordance with National Screening Committee criteria (22).

### Participant experience of T1D screening at the pre-school vaccination visit

It was not clear whether parents fully understood that even if the screening result was negative at the time, that it was still possible that their child could develop positive IAb in the future, despite this information being available. This is consistent with the literature (23). However, all parents stated that they would agree to having their child tested again and they would recommend it to others. The main concern was the perceived volume of blood needed and the time it took to collect. Two alternative approaches to the gold standard method of venous IAb sampling are currently being adopted in the research setting, using capillary whole blood and dried blood spots (DBS). Both methods facilitate screening for GADA/IA-2A/ZnT8A but the DBS method does not usually include the more complex IAA test. As shown in our exploratory study, the new low blood volume LIPS approach (24) allowed screening for all four IAbs in the majority of lower blood volume samples. IAA are often present in the very young child and peak before GADA, IA-2A and ZnT8A. For universal screening, excluding IAA in the first screen would mean that some young children, who have the highest rates of DKA, may be missed. The balance of acceptability and sensitivity will need to be considered, as well as the approach to improving recruitment before further expansion. One approach would be to request lower blood volumes and use LIPS testing or other ultra-low volume technologies (25), rather than RBA, to overcome the concerns raised over blood volume requirements.

### Strengths

Qualitative interviews have been undertaken in antibody positive individuals (26), but to our knowledge have not been undertaken in the general population undergoing screening. In this proof-of-concept study, we report the results from qualitative work undertaken to assess parents’ real-life experience of participating in a potential screening programme. In combination with postcard data, this provided an understanding of the experience of parents on the whole study and will inform future study design.

### Study Limitations

Our study is small and likely biased by being limited to two PCPs from a relatively affluent and white demographic. Non-attenders were not invited for semi-structured interviews.

Future studies should focus on including a larger and more diverse population where vaccination uptake is lower and address how to approach individuals who decline vaccination. Lower sample volume techniques are needed for the purposes of acceptability.

## CONCLUSIONS

This proof-of-concept study shows the potential for screening children for T1D from the general population when aligned to routine pre-school vaccination. Qualitative results indicate general acceptability of the test when taken at the time of the vaccination visit by parents and will inform future expansion.

## Supporting information

Supplemental Material - Participant Information Sheet

## Data Availability

All data produced in the present study are available upon reasonable request to the authors.

## ACKNOWLEDGEMENTS

We would like to thank Dr Eleri Shellens, who provided PPI input into the study, University of Bristol AWAF team members, Rachel Aitken, Anna Long, Kyla Chandler and Olivia Pearce for providing capillary collection kits, sample processing and testing, and the primary care practices in Witney and Bicester who supported the study.

