## Supplemental Materials for "General population screening for type 1 diabetes using islet autoantibodies at the preschool vaccination visit: a proof-of-concept study (the T1Early study)"

**Supplemental Methods**

Supplemental Methods 1: Participant information sheet.

Supplemental Methods 2. Blood collection kits

**Supplemental Tables**

Supplemental Table 1. Eligibility Criteria

Supplemental Table 2. Interview schedule

Supplemental Table 3. Questions asked on Postcards (2A) to participants, and (2B) to non-participants.

**Supplemental Results**

Supplemental Table 4. Self-reported ethnicity and family history of T1D collected using postcard questionnaires

**Supplemental Methods**

**Supplemental Methods 1. Participation information sheet**

**Supplemental Methods 2. Blood collection kits**

Sarstedt safety lancet 1.5mm blade, 1.6mm penetration and Sarstedt Microvette® serum gel capillary tubes with clotting activator (Sarstedt Inc., Newton, NC, USA).

**Supplemental Tables**

**Supplemental Table 1. Eligibility Criteria**

**Inclusion Criteria**

Parent/carer who is willing and able to give informed consent for participation in the study

A well child scheduled to attend a pre-school booster vaccination

Aged ≤ 5 years

**Exclusion Criteria**

Stage 3 T1D

Known coagulopathy or bleeding disorder

Parent/carer has insufficient understanding of written and verbal English

**Supplemental Table 2. Interview Schedule**

| **Initial questions** | |
| --- | --- |
| **1.** | I wonder if you can tell me about your children, their ages, and the age of the child who took part in the recent vaccination/screening programme? |
| **2.** | Please can you now talk me through generally your experience of taking children to be vaccinated? (***Probe as necessary, what made you feel like that? Why?)***  a. Was this the first vaccination experience for this child or had they had previous vaccinations as a baby?  b. What are you general opinions on vaccination/screening?  c. Did your child/children have the heel prick test as baby? What did you think about this at the time?  d. What is your child generally like when you visit the GP/primary care surgery?  e. What were they like with previous vaccinations/injections for anything?  f. Have they been previously unwell which has required them to have a stay in hospital? |
| **3.** | You may remember that the study you took part in was to screen for the antibodies that appear in our blood and show if we are likely to develop Type 1 diabetes.  a. Did you have any knowledge of T1D before taking part in the study?  b. Do you have any family members with T1D?  c. Did you have any previous experience of diabetes?  d. What did you think about T1D? Why? |
| **Prior to vaccination/screening** | |
| **4.** | So I now want you to try and remember what happened when you were offered the vaccination/screening for your child.  a. Can you remember who contacted you?  b. What information were you given? How did you feel?  c. Were you given the opportunity to discuss it with other people?  i)Who did you discuss it with? Family member? Neighbour? Friend?  d. Did you look for further information yourself, independently?  i)Where did you look? What did you find out? Did this influence your decision to take part at all? *(Probe if they needed further information to take part, what sort of things did they want to know?)* |
| **5.** | Overall, what did you think about the opportunity to have your child screened for T1D?  a. Had you heard about T1D prior to the vaccination invitation? What did you understand about the condition prior to the vaccination day?  b. Were you worried or anxious about having your child screened? Why?  c. Did you have any other concerns? What were they? |
| **6.** | Overall why did you decide to have your child screened? |
| **Day of screening** | |
| **7.** | Please could you talk me through exactly what happened, step by step, on the day of the vaccination/screening? ***(Remember any probing questions...How did that make you feel? Why?)***  a. Was it you who took the child?  b. How was your child, happy? Worried? Upset?  c. Who administered the test/vaccine?  d. Did your child have the screening (finger prick blood taken on the day)?  e. Or did you take a kit to do the test at home?  f. How did you feel the vaccination/screening went? Why?  g. What about the person delivering the vaccine/screening? How were they? What did you feel at the time?  h. What could have been done differently?  i. How was your child afterwards? If upset, how long before they were back to their normal selves?  j. What do you think the benefits might be for having your child screened?  k. Have you discussed the screening test with others? Who? Has this changed your opinion in any way? |
| **Whilst waiting for the results** | |
| **8.** | Have you had the results of the screening test yet?  a. Do you know when they are likely to arrive? And how they will arrive? |
| **9.** | Can you describe how you have felt whilst waiting for the results?  a. Have you been worried or not worried at all? If worried, what would have helped reassure you, if anything? Why? Have you sought reassurance from a health professional?  b. Have you sought further information, independently e.g. Google?  c. Overall would you describe the experience as positive or negative? Why?  d. Would you recommend it to other parents? Why? |
| **10.** | What would you think if the vaccination/screening programme was introduced throughout the UK? |
| **11.** | Would you have liked any other information? |
| **12.** | What could have been done differently? |
| **13.** | If made available, say as part of the schools’ vaccination programme, would you have your child screened again? Say when they were 12 or 13? Why? |

**Supplemental Table 3. Questions asked on Postcards (2A) to participants, and (2B) to non-participants.**

|  | **2A - Participants** | **2B – Non-participants** |
| --- | --- | --- |
| **1.** | Tell us about your experience of taking part in T1 Early | Could you tell us why you decided not to take part in the T1 Early Study? |
| **2.** | How did your child find the finger prick blood test? | Is there anything else you would have liked to have known about the study, so that we can improve this in the future? |
| **3.** | Did you complete this card:   - At your child's vaccination visit? - After your child's vaccination visit? | What were the advantages/disadvantages of receiving information electronically |
| **4.** | Why did you decide to take part in the T1 EARLY study? | What is your ethnic group? |
| **5.** | Is there anything else you would have liked to have known before taking part, so that we can improve this in the future? | Does anyone in your immediate family have type 1 diabetes? If yes, who? |
| **6.** | What were the advantages/disadvantages of receiving information electronically/doing remote consent? |  |

**Supplemental Results**

**Supplemental Table 4:** Self-reported ethnicity and family history of T1D collected using postcard questionnaires

|  | **Participants**  **n=29** | **Non-participants**  **n=3** |
| --- | --- | --- |
| **Ethnicity** |  |  |
| English/Welsh/Scottish/Northern Irish/British | 22 | 2 |
| Any other White background, please describe | 1 | 0 |
| Any other Mixed/Multiple ethnic background, please describe | 1 | 0 |
| Indian | 1 | 0 |
| Chinese | 1 | 0 |
| Not stated | 0 | 1 |
| **Family history of T1D** | 3* | 0 |

*1 unknown
