## Supplemental Material - Participant Information Sheet for "General population screening for type 1 diabetes using islet autoantibodies at the preschool vaccination visit: a proof-of-concept study (the T1Early study)"

### T1 Early Study: Establishing the feasibility of antibody screening in primary care at the time of the pre-school vaccination, to identify children with early-onset type 1 diabetes

#### Participant Information Sheet

Your child is invited to take part in our research study, if you wish - it's entirely voluntary.

**When a child is first diagnosed with Type 1 Diabetes (T1D), they can already be very unwell - Help us develop an early detection programme**

To help prevent children from becoming very ill at diagnosis, we have developed an early detection programme. It's a simple fingerprick blood test that is done in seconds by a medical professional - or even you!

The blood is then tested for the early stages of T1D and the results are sent to you within eight weeks.

This test could be life-changing, preventing avoidable illness and family stress. But first, we need to do this study, to understand your experience of taking part in order to guide us in the development of a national diabetes testing programme for all children.

Involving 60 pre-school children across two GP practices in the Thames Valley area, we hope this may be the first step in a national programme, helping to protect all children. If you have any queries after reading this leaflet and before making your decision, please get in touch - our contact details are at the end.

### Your Questions Answered

#### Why is early detection of T1D important?

- T1D destroys the cells needed for the body to make insulin, which means your blood sugar levels rise. This makes children very unwell if not treated quickly. Most children with T1D do not have a family history of diabetes, which could leave your family unprepared.

- The classic symptoms of T1D are drinking and weeing more than usual, being unusually tired and losing weight, but can be non-specific.

- T1D can be diagnosed through markers in the blood years before symptoms appear - these markers are called diabetes antibodies.

- Detecting two or more diabetes antibodies in the blood tells us that T1D is going to happen; there is an 80-90% chance of your child needing insulin before the age of 18 years. We can then prepare you and your child for that diagnosis.

#### What are the advantages of taking part in the study?

- There may be no direct benefit to you or your child, as the chances of a positive test result are low as we will only be testing 60 children for this study (1 in 350 children are diagnosed with type 1 diabetes).

- If we do find a positive result, it allows you and your child to come to terms with the diagnosis and learn about diabetes and its treatment while your child remains well.

- With your permission, we can notify your child's GP.

- You and your doctors can then act quickly if symptoms appear, avoiding the risk of serious illness with DKA.

- There may be future opportunities for children at risk of developing T1D to take part in other research, with medications to delay or prevent diabetes developing.

#### What are the possible disadvantages of taking part in the study?

- The finger prick blood test may hurt a little but it will be over quickly. There may be a small amount of redness and temporary soreness. Keep a treat handy to reward them for being brave!

- If you learn that your child is at risk of developing T1D in the future, this could make you worried. To help, we will invite you to meet a diabetes health professional who will explain all about T1D. If you become very anxious, we can put you in touch with a hospital psychologist.

#### What happens if I decide my child can take part?

- You will be asked to complete a consent form; this can be done prior to your visit on a telephone or video call with the study nurse, or if you prefer, in-person with the study nurse at your visit.

- The nurse will, ask you a few questions about your child and their medical and family history, and then collect a few drops of blood from your child's finger using a small finger pricker and collection tube. This sample is then sent for diabetes antibody testing.

- You'll then be given a postcard with a few short questions about your child's experience of the testing and your experience of the test. Please complete and return the prepaid postcard to us.

#### What if I want my child to have the test but they don't want it done at the time of the pre-school vaccination?

- If preferred, you can arrange an appointment for another day, or we will give you a kit with instructions on collecting the sample yourself at home to post back to the laboratory.

#### Test Results - what do they mean?

We're testing for four different diabetes antibodies to detect your child's risk of developing T1D in the future. We will let you have the test results by letter, email or telephone in about eight weeks.

- Lower risk: This is when no diabetes antibodies are found, being a negative result. This does not guarantee that your child will never get T1D as antibodies can still develop later.

- Low risk: If one diabetes antibody is found, this is a positive result but does not mean your child will develop T1D. There may be a slight increased risk. A re-test will be offered.

- High risk: If two or more diabetes antibodies are found, this shows that your child is already in the early stage of T1D. Your child's body should still control its blood sugar level now, but insulin may be needed in the future. We will invite you for a re- test to confirm the results and discuss them with you.

#### Do I have to take part? Can I change my mind?

- You do not have to take part; this is completely voluntary, and your decision either way will not affect your child's pre-school vaccination visit or any future medical care in any way.

- You can agree and then change your mind and withdraw without giving a reason. This will not affect your child's medical care.

- If you and your child withdraw from the study, we will keep the information about your child that we have already obtained. If you withdraw before your child's sample has been analysed then we will discard the sample, otherwise the results will be retained by the lab, but not fed back to you or your GP.

- Since this is a feasibility study, we would be interested to understand why you may decide not to get your child tested and would appreciate you answering a few questions. These will be given to you at your child's pre-school vaccination or sent in the post from your GP. Please then return the prepaid postcard to us - your answers will be valuable when designing future studies.

#### Participation in future research

We hope to undertake further research (interviews/questionnaires) with families who take part in the study, and also those that chose not to take part. Agreeing to being contacted does not oblige you to take part.

#### How will information be kept confidential?

- All information collected about you and your child during the study will be kept strictly confidential. Your child's sample and any information recorded will be assigned a study code; the identity of the volunteers will not be available to anyone in the research team who will use the samples.
- Your child's anonymised information will be held in a secure database located at the Diabetes and Inflammation Laboratory, Wellcome Centre for Human Genetics, University of Oxford.
- Responsible members of the University of Oxford, relevant NHS Trusts or Regulatory Agencies may be given access to data for monitoring and/or audit of the study to ensure we are complying with regulations.

#### What will happen to my child's data?

- Data protection regulation requires that we state the legal basis for processing information about you. In the case of research, this is 'a task in the public interest.' The University of Oxford is the sponsor for this study, based in the United Kingdom, is the data controller and is responsible for looking after your information and using it properly.

- We will be using information from you and your child in order to undertake this study, and will use the minimum personally identifiably information possible. We will keep identifiable information about you and your child for three years after the last participant has completed the study. This excludes any research documents with personal information, such as consent forms, which will be held securely at the University of Oxford after the end of the study, until the youngest participant has reached 21 years of age.

- If you agree to be contacted about other research, we will hold your contact details securely, separately from the study data. You can request to be removed from this register at any time.

- The GP surgery will use your child's name, NHS number, home address and contact details to contact you and your child about the research study, to ensure that relevant information is recorded for your child's care, and to oversee the quality of the study. They will keep identifiable information about your child from this study in keeping with their policies for retention of medical notes.

- Data protection regulation provides you with control over your personal data and how it is used. When you agree to your information being used in research, however, some of those rights may be limited in order for the research to be reliable and accurate. Further information about your rights with respect to your personal data is available at: https://compliance.web.ox.ac.uk/individual-rights

#### What will happen to my child's samples?

- Your child's blood sample will be stored securely and analysed in a laboratory in the UK.

- Once analysed, any remaining sample will be destroyed.

#### Who has reviewed this study?

- All research in the NHS is looked at by an independent group of people, called a Research Ethics Committee, to protect participants' interests. This study has been reviewed and given a favourable opinion by the North West Preston Research Ethics Committee.

#### Who is organising and funding the research?

- This study is sponsored by the University of Oxford and is funded by the Wellcome.

#### What if there is a problem?

- If you wish to complain about any aspect of the way in which you have been approached or treated during the course of this study, you should contact the study team either on 07765 932065 or by emailing, or you may contact the University of Oxford Research Governance, Ethics and Assurance Team (RGEA) office on 01865 61648 or email the director of RGEA at.

- The University of Oxford, as Sponsor, has appropriate insurance in place in the unlikely event that you suffer any harm as a direct consequence of your participation in this study.

#### If you are interested in taking part, or would like further information, please contact:

- Jane Bowen-Morris, Lead Research Nurse, T1 Early study,
